# Survival after deceased-donor kidney transplantation versus continued waiting among candidates with kidney failure attributed to renal cell carcinoma

**DOI:** 10.64898/2026.11.21.26368667

**Authors:** Junho Song, Esther Kim, Amar Kassim Amar, Julian Chavarriaga Soto, Jay D. Raman, Jaime Herrera-Caceres

## Abstract

**Background:** Among transplant candidates with kidney failure attributed to renal cell carcinoma (RCC), we investigated whether deceased-donor kidney transplantation was associated with longer survival than continued waiting and whether the association varied by kidney donor profile index (KDPI).

**Methods:** We studied adult kidney-alone candidates listed in 2005-2025 in the Organ Procurement and Transplantation Network registry (follow-up through March 31, 2026) whose coded cause of kidney failure was RCC, versus all other candidates. Time-dependent Cox models compared transplantation with waiting. Calendar-aligned sequential trials with propensity overlap weighting estimated survival after transplantation with a KDPI ≤85% kidney versus continued waiting.

**Results:** Of 599,887 candidates, 1,542 had RCC-attributed kidney failure (614 deaths). Of these, 597 received a KDPI ≤85% kidney and 38 received a KDPI >85% kidney. A KDPI ≤85% transplant was associated with lower mortality than waiting (hazard ratio [HR] 0.51, 95% confidence interval [CI] 0.42 to 0.62). The HR was 0.53 in other candidates (interaction P = 0.86). Five-year survival was 83.1% after transplantation and 67.0% with continued waiting (difference, 16.0 percentage points, 95% CI 11.8 to 20.1). The restricted mean survival time difference was 5.8 months (95% CI 4.4 to 7.4). For KDPI >85%, the HR was 1.65 (95% CI 0.87 to 3.13).

**Conclusions:** Transplantation with a KDPI ≤85% kidney was associated with about 16 additional survivors per 100 RCC candidates at 5 years. The estimate for KDPI >85% was imprecise. Tumor stage and cancer-free interval were unavailable.

## Introduction

Renal cell carcinoma (RCC) and its treatment can compromise kidney function. Radical nephrectomy is an independent risk factor for incident chronic kidney disease after surgery for small renal tumors, and many patients have impaired kidney function before surgery.^1^ When surgery is superimposed on chronic kidney disease, rates of progressive kidney function decline and death are higher.^2^ Current guidance therefore emphasizes preservation of kidney function alongside oncologic control.^3^ Despite nephron-sparing strategies, some patients ultimately progress to kidney failure after radical or bilateral nephrectomy, nephrectomy of a solitary kidney, or progression of underlying kidney disease and are subsequently referred for kidney transplantation. Urologists often follow these patients through oncologic surveillance and are therefore well positioned to counsel them regarding transplant referral and candidacy.

For most patients with kidney failure, transplantation is associated with longer survival than continued dialysis while awaiting transplantation.^4^ However, patients treated for RCC pose a specific challenge. Kidney transplant recipients have a higher incidence of RCC than the general population.^5^ Transplantation is generally deferred until a stage-and grade-specific cancer-free interval has elapsed.^6,7^ A systematic review found similar recurrence and survival after treated RCC among transplant recipients and dialysis patients.^8^ Once such a patient is listed, however, the question shifts from candidacy to timing: when a deceased-donor kidney is offered, should the patient accept it or continue waiting for a better offer?

The kidney donor profile index (KDPI), derived from the kidney donor risk index,^9^ summarizes donor quality on a scale from 0% to 100% and has guided kidney allocation in the United States since implementation of the Kidney Allocation System (KAS) in December 2014.^10^ In the general waitlisted population, even kidneys from higher-risk donors are associated with long-term survival benefit compared with waiting for a better offer.^11,12^ However, candidates listed after RCC are older and face competing oncologic risks that could attenuate this benefit. A previous registry analysis reported better outcomes among transplanted than still-waiting candidates with kidney failure attributed to RCC,^13^ but did not model transplantation as a time-varying exposure, examine donor quality, or compare the association with that observed in other candidates.

Consequently, whether the survival benefit of deceased-donor kidney transplantation, and its dependence on donor quality, is preserved among candidates with RCC remains uncertain.

We therefore investigated, among transplant candidates whose kidney failure was attributed to RCC, whether receiving a deceased-donor kidney was associated with longer survival than continued waiting, whether this association varied by KDPI, and whether it differed from that observed in other candidates. We report relative estimates from time-dependent Cox models and absolute estimates from calendar-aligned sequential trials indexed on the date of transplantation.

## Materials and Methods

### Data Source and Study Population

This retrospective cohort study used the Organ Procurement and Transplantation Network (OPTN)/United Network for Organ Sharing Standard Transplant Analysis and Research files (release with data through March 31, 2026). We included adult candidates first listed for a kidney-alone transplant from 2005 to 2025. Of 600,005 candidates, 118 with a recorded death date before listing (110) or before a recorded transplant (8) were excluded, leaving 599,887 (Figure 1). Candidates whose coded cause of kidney failure at listing was RCC (diagnosis code 3021) formed the study group; all other candidates formed the non-RCC reference cohort, used to assess whether the association between transplantation and mortality differed by RCC status. This study used de-identified registry data from the United Network for Organ Sharing (UNOS). Institutional review board approval and informed consent were not required for this secondary analysis of de-identified data. Reporting follows the STROBE Statement.^14^

**Figure 1.**
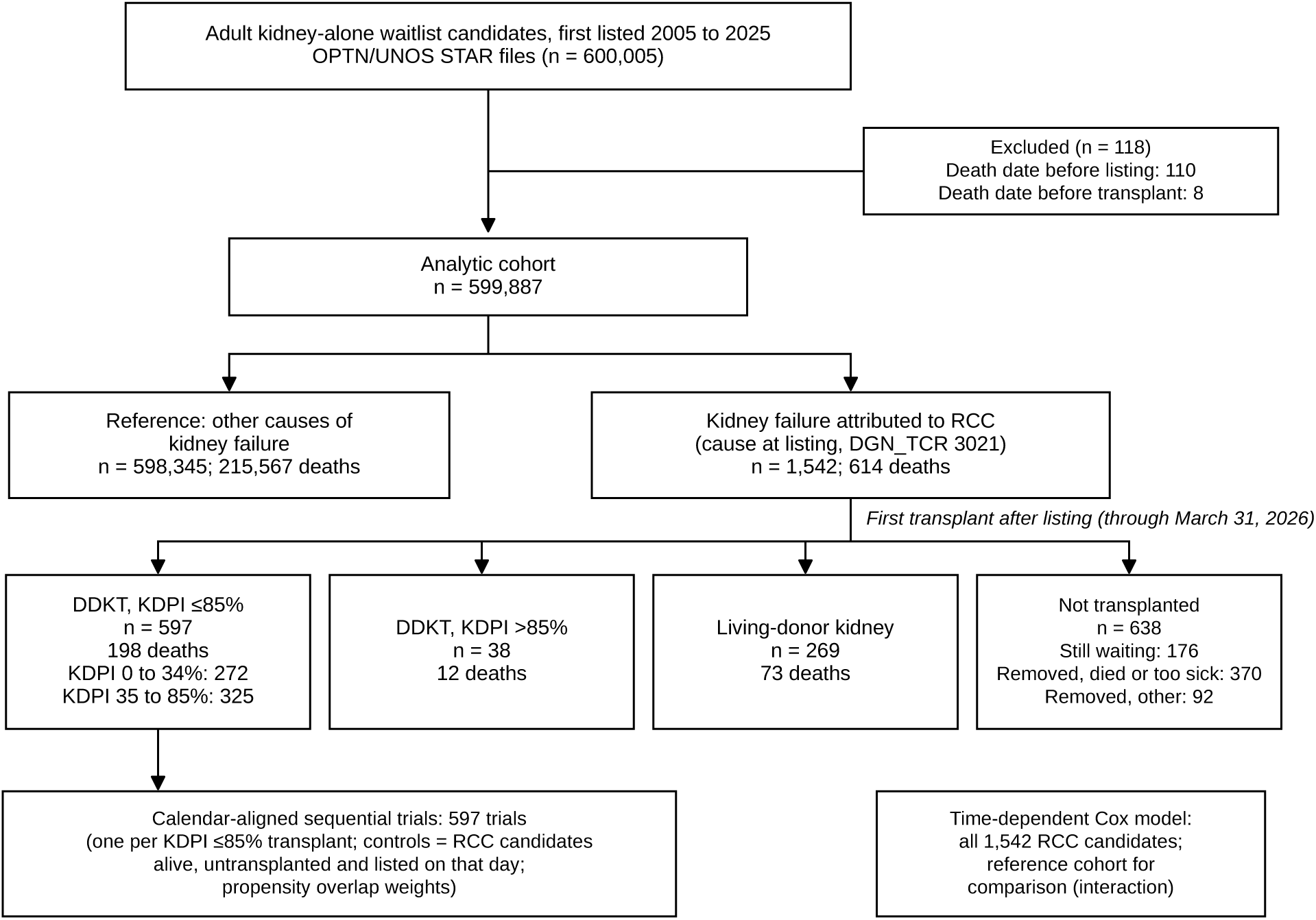
Cohort flow. Adult kidney-alone candidates first listed from 2005 to 2025 in the OPTN registry, split into the reference cohort and candidates with kidney failure attributed to RCC, and the RCC candidates by first transplant after listing through March 31, 2026. Counts of deaths are deaths from any cause during follow-up. DDKT, deceased-donor kidney transplantation; DGN_TCR, registry code for the cause of kidney failure at listing; KDPI, kidney donor profile index; OPTN, Organ Procurement and Transplantation Network; RCC, renal cell carcinoma; STAR, Standard Transplant Analysis and Research; UNOS, United Network for Organ Sharing.

### Exposure States and Outcome

Time zero was the date of first listing. Each candidate contributed time in the waiting state until the first transplant after listing, if any, and thereafter in the state defined by that transplant: deceased-donor kidney transplantation (DDKT) with KDPI ≤85% (subdivided into 0% to 34% and 35% to 85%), DDKT with KDPI >85%, or living-donor kidney transplantation (LDKT). KDPI applies only to deceased-donor kidneys; LDKT was modeled separately and was not assigned a KDPI. The outcome was death from any cause, taken from the patient-level composite death date across waitlist and transplant records, so that follow-up continued after removal from the list and after transplantation; follow-up was censored on March 31, 2026. Deaths on the day of transplantation were attributed to the transplant state. For the primary RCC analysis, the reference state was time spent waiting among RCC candidates. This included time before a later transplant as well as waiting time among candidates who never received a transplant.

### Covariates

Models adjusted for characteristics at listing: age group (18 to 44, 45 to 54, 55 to 64, and ≥65 years), sex, race and ethnicity, diabetes, dialysis duration at listing (preemptive, <1, 1 to 3, and ≥3 years), calculated panel-reactive antibody (0%, 1% to 79%, ≥80%, or unknown), ABO blood group, primary payer, functional status, prior kidney transplant, and listing era (before KAS; KAS from December 4, 2014; acuity circles from March 15, 2021). Missing values of categorical covariates were coded as an explicit unknown level; body mass index (missing for 3,727 reference and 6 RCC candidates) was described but not modeled. Inactive status at listing was not a model covariate but defined a sensitivity analysis.

### Time-Dependent Cox Models

Hazard ratios (HRs) for each transplant state versus waiting were estimated with Cox proportional hazards models in counting-process form, with transplant state as a time-varying exposure to avoid immortal time bias.^4,15^ Ties were handled with the Breslow method, and 95% confidence intervals (CIs) used cluster-robust standard errors by listing center. The primary model separated KDPI ≤85%, KDPI >85%, and LDKT; secondary models used KDPI tiers, any DDKT, a 50% KDPI cut point, and continuous KDPI among DDKT recipients. To describe how the association changes after transplantation, we split post-transplant time at 6 months and fitted a restricted cubic spline in log time since DDKT (knots at 7, 30, 120, 365, and 1,826 days), from which we derived the day of equal instantaneous risk and the day of equal cumulative survival with bootstrap CIs. The same models were fitted in the full reference cohort (the spline model in a random 25% sample). A pooled model with transplant state by RCC interaction terms, and baseline hazards stratified by RCC, tested whether HRs differed (ratio of HRs, RCC versus reference).

### Calendar-Aligned Sequential Trials

To express the primary comparison in absolute terms, we emulated a sequence of target trials.^16^ Each RCC candidate who received a KDPI ≤85% kidney defined one trial whose time zero was the calendar date of that transplant. Individual offers and refusals were unavailable. Controls were RCC candidates who were alive, listed, and not yet transplanted on that date, and were followed regardless of later transplantation. Up to 50 controls were sampled per trial, weighted by the risk-set size divided by the number sampled. A propensity model included the listing covariates, time since listing at trial entry, and allocation era at trial entry, and overlap weights were applied.^17^ Weighted Kaplan-Meier curves gave survival and between-group survival differences at 1, 3, and 5 years, as well as the difference in restricted mean survival time (RMST) over 5 years. CIs came from a candidate-level bootstrap (B = 300) that rebuilt the trials and refitted the propensity model in each replicate. The number of candidates who would need to accept a kidney for one additional 5-year survivor was the reciprocal of the 5-year survival difference, with CI limits obtained by inverting those of the difference.

### Sensitivity and Additional Analyses

Sensitivity analyses of the primary HR were restricted to KAS-era listings, excluded candidates inactive at listing, were restricted to age ≥60 years, and censored follow-up at LDKT, at removal from the list for reasons other than death or transplantation, or at multiorgan transplantation. Waiting-time-aligned sequential trials, in which controls were aligned on time since listing rather than calendar date, served as an alternative design.

A prespecified exploratory analysis estimated the primary HR within age strata (<65 and ≥65 years, with linear age in place of age group) and compared stratum estimates with a z test. The E-value for the primary HR was computed with the common-outcome conversion.^18^ Cancer deaths and graft failure among deceased-donor recipients whose diagnosis at transplant was RCC are described in Table S9.

### Software and Verification

The original analysis used Python 3.13 with lifelines, statsmodels and scikit-learn. All headline estimates were recomputed independently in Python 3.10 with NumPy and pandas from the analytic file, including the full reference cohort without sampling and the sequential trials with all eligible controls; all 171 verification records agreed within prespecified tolerances (SDC, Materials and Methods, and Table S10). Two-sided P < 0.05 was considered statistically significant.

ChatGPT (OpenAI) was used for copyediting and manuscript formatting. The authors are responsible for the manuscript content.

## Results

### Cohort

Of 599,887 candidates, 1,542 (0.3%) had kidney failure attributed to RCC (Figure 1). Compared with the reference cohort, RCC candidates were older (median age 63 vs 55 years; standardized mean difference [SMD] 0.78), more often male (75.3% vs 61.6%) and White (64.0% vs 43.4%), and less often had diabetes (22.4% vs 46.9%; Table 1; Table S1). During 10,209 person-years (median potential follow-up 9.3 years), 614 RCC candidates (39.8%) died. A KDPI ≤85% kidney was received by 597 candidates (272 KDPI 0% to 34%, 325 KDPI 35% to 85%) at a median of 541 days after listing, a KDPI >85% kidney by 38, and a living-donor kidney by 269. Of the 638 candidates who were not transplanted, 176 were still waiting at the end of follow-up, 370 had been removed because of death or deteriorating health, and 92 had been removed for other reasons. Overall, 331 deaths occurred in the waiting state and 198 after a KDPI ≤85% transplant.

**Table 1.** Characteristics at listing of the reference cohort and of candidates with kidney failure attributed to RCC.

| **Characteristic** | **Reference** | **RCC** | **SMD, RCC vs ref** |
|---|---|---|---|
| Candidates, n | 598,345 | 1,542 |  |
| Age, years, median (IQR) | 55.0 (44.0 to 63.0) | 63.0 (56.0 to 69.0) | 0.78 |
| Dialysis duration at listing, years, median (IQR) | 0.5 (0.0 to 1.6) | 0.8 (0.0 to 2.4) | 0.15 |
| Body mass index, kg/m<sup>2</sup>, median (IQR) | 28.6 (24.8 to 32.9) | 29.3 (25.6 to 32.9) | 0.07 |
| Missing, n | 3,727 | 6 |  |
| Age group, years, n (%) |  |  | 0.58 |
| 18 to 44 | 156,408 (26.1) | 87 (5.6) |  |
| 45 to 54 | 141,605 (23.7) | 231 (15.0) |  |
| 55 to 64 | 179,460 (30.0) | 544 (35.3) |  |
| ≥65 | 120,872 (20.2) | 680 (44.1) |  |
| Male sex, n (%) | 368,653 (61.6) | 1,161 (75.3) | 0.30 |
| Race and ethnicity, n (%) |  |  | 0.42 |
| White | 259,597 (43.4) | 987 (64.0) |  |
| Black | 169,034 (28.3) | 375 (24.3) |  |
| Hispanic | 114,532 (19.1) | 133 (8.6) |  |
| Asian | 41,982 (7.0) | 34 (2.2) |  |
| Other | 13,200 (2.2) | 13 (0.8) |  |
| Diabetes, n (%) | 280,583 (46.9) | 346 (22.4) | −0.53 |
| Dialysis duration at listing, n (%) |  |  | 0.19 |
| Preemptive | 212,809 (35.6) | 528 (34.2) |  |
| <1 y | 166,621 (27.8) | 305 (19.8) |  |
| 1 to 3 y | 146,706 (24.5) | 436 (28.3) |  |
| ≥3 y | 72,209 (12.1) | 273 (17.7) |  |
| Calculated panel-reactive antibody, %, n (%) |  |  | 0.06 |
| 0 | 382,482 (63.9) | 960 (62.3) |  |
| 1 to 79 | 43,051 (7.2) | 103 (6.7) |  |
| ≥80 | 12,113 (2.0) | 21 (1.4) |  |
| Unknown | 160,699 (26.9) | 458 (29.7) |  |
| ABO blood group, n (%) |  |  | 0.08 |
| O | 289,411 (48.4) | 729 (47.3) |  |
| A | 197,764 (33.1) | 565 (36.6) |  |
| B | 87,389 (14.6) | 190 (12.3) |  |
| AB | 23,781 (4.0) | 58 (3.8) |  |
| Primary payer, n (%) |  |  | 0.16 |
| Private | 269,956 (45.1) | 574 (37.2) |  |
| Public | 324,929 (54.3) | 957 (62.1) |  |
| Other or unknown | 3,460 (0.6) | 11 (0.7) |  |
| Functional status, n (%) |  |  | 0.04 |
| Normal or minimal limitation | 375,501 (62.8) | 1,000 (64.9) |  |
| Impaired | 198,316 (33.1) | 490 (31.8) |  |
| Unknown | 24,528 (4.1) | 52 (3.4) |  |
| Prior kidney transplant, n (%) | 14,008 (2.3) | 6 (0.4) | −0.17 |
| Inactive status at listing, n (%) | 170,266 (28.5) | 537 (34.8) | 0.14 |
| Listing era, n (%) |  |  | 0.08 |
| Before KAS (before Dec 4, 2014) | 255,270 (42.7) | 656 (42.5) |  |
| KAS (Dec 4, 2014 to Mar 14, 2021) | 175,598 (29.3) | 397 (25.7) |  |
| Acuity circles (from Mar 15, 2021) | 167,477 (28.0) | 489 (31.7) |  |
| **Follow-up** |  |  |  |
| Deaths, n (%) | 215,567 (36.0) | 614 (39.8) |  |
| Person-years | 4,346,865 | 10,209 |  |
| Median potential follow-up, y | 9.7 | 9.3 |  |

Values are n (%) or median (IQR); characteristics of RCC candidates by eventual transplant status are given in Table S1. SMD is RCC versus reference; for categorical variables with more than two levels, the largest absolute level SMD is shown. Potential follow-up is the time from listing to the data end date. IQR, interquartile range; KAS, Kidney Allocation System; RCC, renal cell carcinoma; ref, reference cohort; SMD, standardized mean difference.

### Transplant State and Mortality

Compared with continued waiting, DDKT with a KDPI ≤85% kidney was associated with lower mortality among RCC candidates (HR 0.51, 95% CI 0.42 to 0.62; Table 2, Figure 2). Associations were similar for KDPI 0% to 34% (HR 0.45, 95% CI 0.35 to 0.59) and 35% to 85% (HR 0.57, 95% CI 0.46 to 0.71), and LDKT, analyzed separately from the KDPI categories, was associated with an HR of 0.39 (0.30 to 0.50). Any DDKT was associated with an HR of 0.54 (0.45 to 0.65); HRs were 0.49 (0.39 to 0.62) for KDPI below 50% and 0.64 (0.50 to 0.82) for KDPI 50% or higher, and among DDKT recipients each 10-point higher KDPI corresponded to an HR of 1.08 (1.01 to 1.16) (Tables S2 and S3). For KDPI >85% kidneys (38 recipients, 12 deaths), no survival benefit was demonstrated (HR 1.65, 95% CI 0.87 to 3.13).

**Figure 2.**
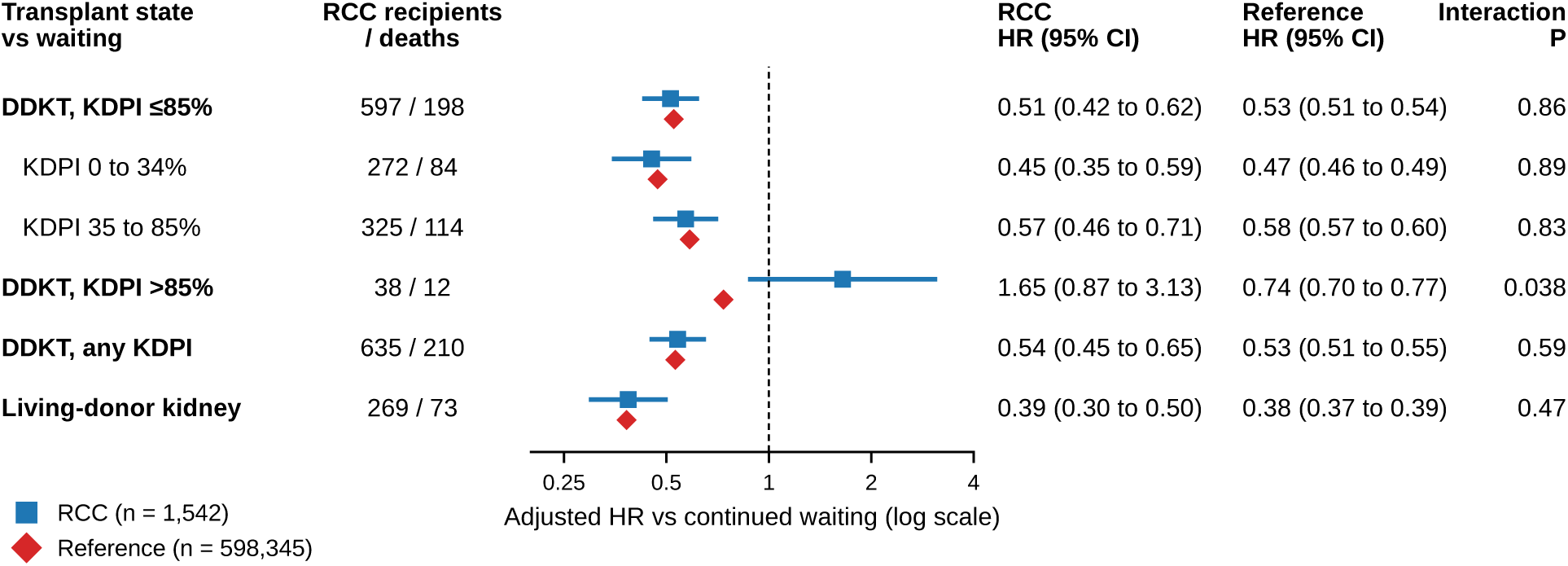
Adjusted hazard ratios for death for each transplant state versus continued waiting. Blue squares, RCC candidates; red diamonds, reference cohort; horizontal lines, 95% CIs (log scale). Estimates are from time-dependent Cox models adjusted for characteristics at listing; interaction P compares the RCC and reference HRs in a pooled model. CI, confidence interval; DDKT, deceased-donor kidney transplantation; HR, hazard ratio; KDPI, kidney donor profile index; RCC, renal cell carcinoma.

**Table 2.**
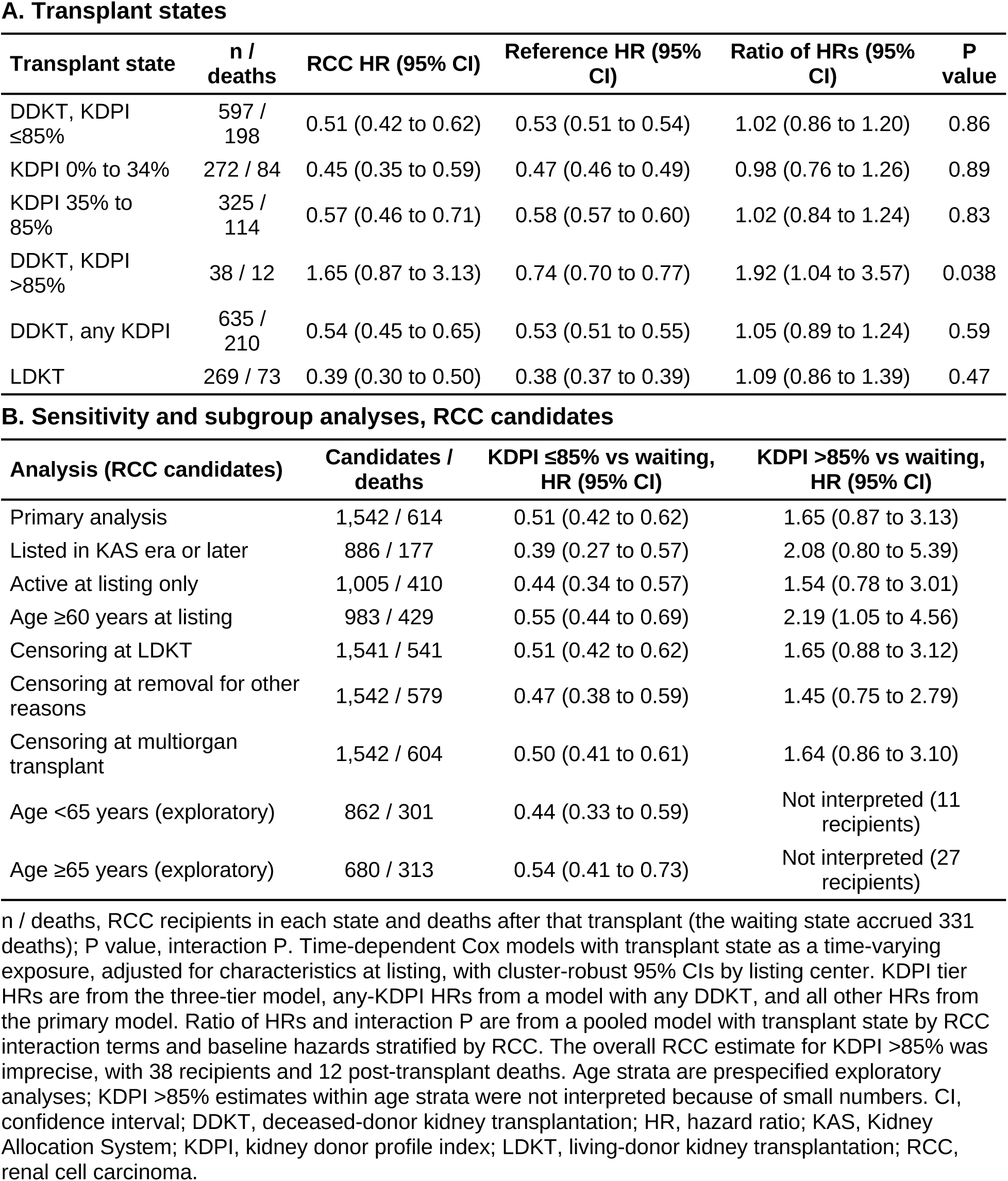
Adjusted hazard ratios for death for each transplant state versus continued waiting, RCC and reference cohorts, and sensitivity analyses in RCC candidates.

| A. Transplant states |  |  |  |  |  |
|---|---|---|---|---|---|
| Transplant state | n / deaths | RCC HR (95% CI) | Reference HR (95% CI) | Ratio of HRs (95% CI) | P value |
| DDKT, KDPI ≤85% | 597 / 198 | 0.51 (0.42 to 0.62) | 0.53 (0.51 to 0.54) | 1.02 (0.86 to 1.20) | 0.86 |
| KDPI 0% to 34% | 272 / 84 | 0.45 (0.35 to 0.59) | 0.47 (0.46 to 0.49) | 0.98 (0.76 to 1.26) | 0.89 |
| KDPI 35% to 85% | 325 / 114 | 0.57 (0.46 to 0.71) | 0.58 (0.57 to 0.60) | 1.02 (0.84 to 1.24) | 0.83 |
| DDKT, KDPI >85% | 38 / 12 | 1.65 (0.87 to 3.13) | 0.74 (0.70 to 0.77) | 1.92 (1.04 to 3.57) | 0.038 |
| DDKT, any KDPI | 635 / 210 | 0.54 (0.45 to 0.65) | 0.53 (0.51 to 0.55) | 1.05 (0.89 to 1.24) | 0.59 |
| LDKT | 269 / 73 | 0.39 (0.30 to 0.50) | 0.38 (0.37 to 0.39) | 1.09 (0.86 to 1.39) | 0.47 |
| **B. Sensitivity and subgroup analyses, RCC candidates** |  |  |  |  |  |
| **Analysis (RCC candidates)** | **Candidates / deaths** | **KDPI ≤85% vs waiting, HR (95% CI)** | **KDPI >85% vs waiting, HR (95% CI)** |  |  |
| Primary analysis | 1,542 / 614 | 0.51 (0.42 to 0.62) | 1.65 (0.87 to 3.13) |  |  |
| Listed in KAS era or later | 886 / 177 | 0.39 (0.27 to 0.57) | 2.08 (0.80 to 5.39) |  |  |
| Active at listing only | 1,005 / 410 | 0.44 (0.34 to 0.57) | 1.54 (0.78 to 3.01) |  |  |
| Age ≥60 years at listing | 983 / 429 | 0.55 (0.44 to 0.69) | 2.19 (1.05 to 4.56) |  |  |
| Censoring at LDKT | 1,541 / 541 | 0.51 (0.42 to 0.62) | 1.65 (0.88 to 3.12) |  |  |
| Censoring at removal for other reasons | 1,542 / 579 | 0.47 (0.38 to 0.59) | 1.45 (0.75 to 2.79) |  |  |
| Censoring at multiorgan transplant | 1,542 / 604 | 0.50 (0.41 to 0.61) | 1.64 (0.86 to 3.10) |  |  |
| Age <65 years (exploratory) | 862 / 301 | 0.44 (0.33 to 0.59) | Not interpreted (11 recipients) |  |  |
| Age ≥65 years (exploratory) | 680 / 313 | 0.54 (0.41 to 0.73) | Not interpreted (27 recipients) |  |  |

n / deaths, RCC recipients in each state and deaths after that transplant (the waiting state accrued 331 deaths); P value, interaction P. Time-dependent Cox models with transplant state as a time-varying exposure, adjusted for characteristics at listing, with cluster-robust 95% CIs by listing center. KDPI tier HRs are from the three-tier model, any-KDPI HRs from a model with any DDKT, and all other HRs from the primary model. Ratio of HRs and interaction P are from a pooled model with transplant state by RCC interaction terms and baseline hazards stratified by RCC. The overall RCC estimate for KDPI >85% was imprecise, with 38 recipients and 12 post-transplant deaths. Age strata are prespecified exploratory analyses; KDPI >85% estimates within age strata were not interpreted because of small numbers. CI, confidence interval; DDKT, deceased-donor kidney transplantation; HR, hazard ratio; KAS, Kidney Allocation System; KDPI, kidney donor profile index; LDKT, living-donor kidney transplantation; RCC, renal cell carcinoma.

For KDPI ≤85% and LDKT, the HRs for RCC candidates closely matched those of the reference cohort, whereas for KDPI >85% they did not. For KDPI ≤85%, the reference HR was 0.53 (0.51 to 0.54) and the ratio of HRs was 1.02 (0.86 to 1.20; P = 0.86); for LDKT, the ratio was 1.09 (0.86 to 1.39; P = 0.47). For KDPI >85%, the reference HR was 0.74 (0.70 to 0.77) and the ratio of HRs was 1.92 (1.04 to 3.57; P = 0.038), based on 38 RCC recipients.

The association changed with time since transplantation (Figure S1). In the first 6 months after a KDPI ≤85% transplant, the HR was 0.83 (95% CI 0.49 to 1.40). The interval includes both lower and higher mortality than with waiting, so the early association was inconclusive. Thereafter, mortality was lower (HR 0.49, 95% CI 0.40 to 0.59). The instantaneous risk of death after DDKT equaled that of waiting at day 29 (95% CI 11 to 107), and cumulative survival became equal at day 240 (72 to 590), similar to the reference cohort (days 33 and 218; Table S4). After a KDPI >85% transplant, early mortality exceeded that of waiting (HR in the first 6 months 4.85, 95% CI 1.93 to 12.21; later 1.22, 95% CI 0.59 to 2.51).

### Absolute Survival After Transplantation Versus Continued Waiting

In 597 calendar-aligned trials, overlap weighting balanced all covariates (maximum absolute SMD 0.42 before and <0.001 after weighting). Survival after receiving a KDPI ≤85% kidney versus continued waiting was 96.9% versus 91.6% at 1 year, 90.0% versus 78.1% at 3 years, and 83.1% (95% CI 79.0 to 86.6) versus 67.0% (64.0 to 70.8) at 5 years (Figure 3). The survival difference was 5.4 percentage points at 1 year (95% CI 3.5 to 7.1) and 11.9 percentage points at 3 years (95% CI 8.8 to 15.3); both intervals excluded zero. The 5-year survival difference was 16.0 percentage points (95% CI 11.8 to 20.1) and the 5-year RMST gain 5.8 months (4.4 to 7.4), corresponding to one additional 5-year survivor for every 6.2 candidates (5.0 to 8.5) who accepted. Of the 597 recipients, 81 died within 5 years of transplantation. With all eligible controls instead of sampled controls, the difference was 16.0 points (11.5 to 20.0; Tables S5 and S6).

**Figure 3.**
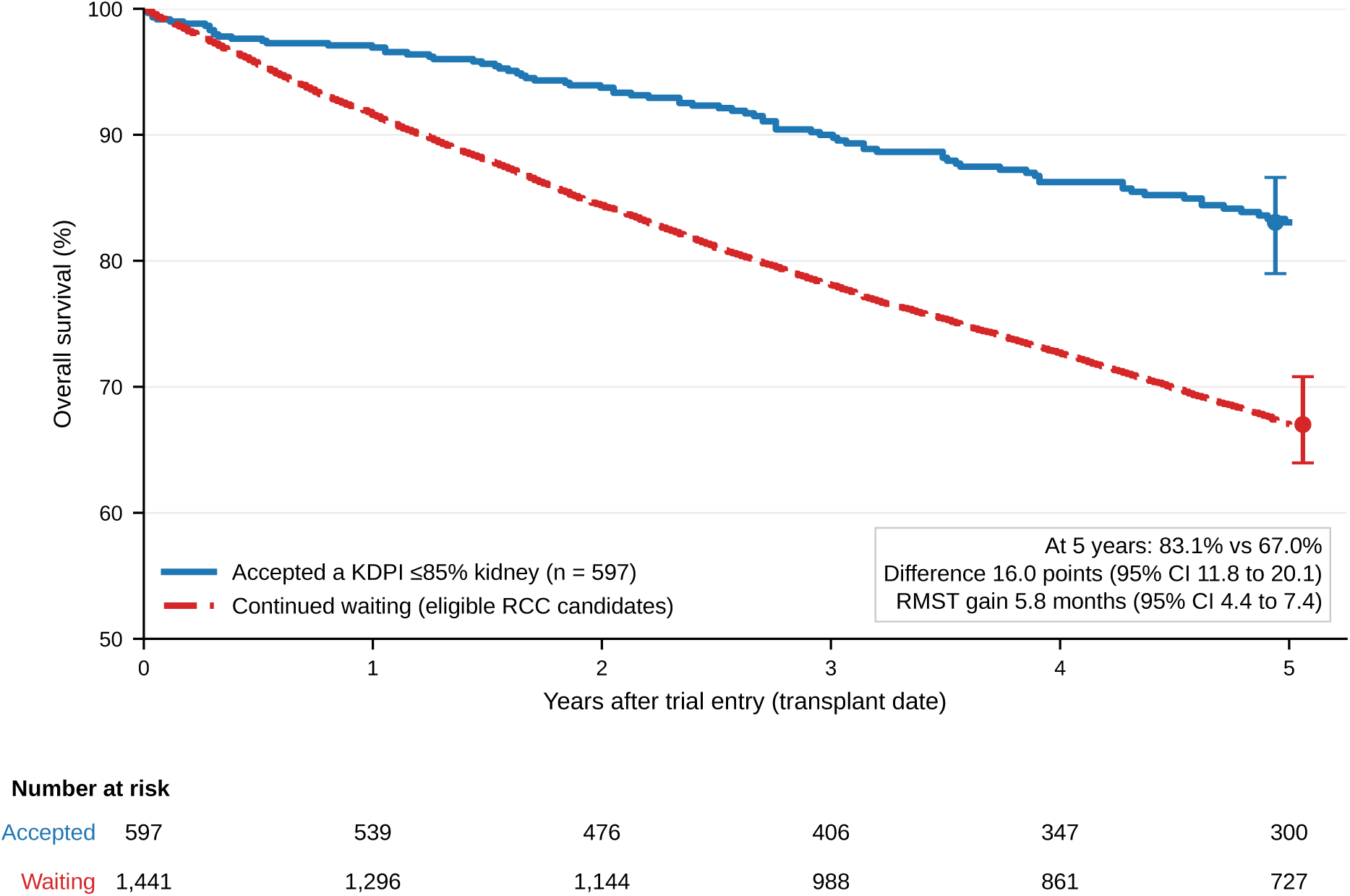
Overlap-weighted survival after receiving a KDPI ≤85% kidney versus continued waiting among RCC candidates, from 597 calendar-aligned sequential trials. Time zero is the calendar date of each transplant; the waiting arm includes candidates who were alive, listed and untransplanted on that date, followed regardless of later transplantation. Whiskers show bootstrap 95% CIs at 5 years. Numbers at risk are recipients (transplant group) and unique candidates (waiting). CI, confidence interval; KDPI, kidney donor profile index; RCC, renal cell carcinoma; RMST, restricted mean survival time.

### Sensitivity Analyses

The primary HR ranged from 0.39 to 0.55 across sensitivity analyses (Table 2; Tables S7 and S8), including KAS-era listings (HR 0.39, 95% CI 0.27 to 0.57) and candidates active at listing (HR 0.44, 95% CI 0.34 to 0.57). Waiting-time-aligned trials gave a 5-year difference of 14.0 points (10.3 to 17.6) and an RMST gain of 4.3 months (3.1 to 5.6). In exploratory age strata, the HR was 0.44 (0.33 to 0.59) below age 65 years and 0.54 (0.41 to 0.73) at 65 years or older (ratio 1.24, P = 0.31). The E-value was 2.54 for the point estimate and 2.12 for the CI limit.

## Discussion

In this national cohort of transplant candidates whose kidney failure was attributed to RCC, receiving a deceased-donor kidney with KDPI ≤85% was associated with about half the mortality of continued waiting, an association similar to that in other candidates. In the calendar-aligned analysis, estimated 5-year survival was 83.1% after transplantation and 67.0% with continued waiting. This corresponds to about 16 additional survivors per 100 candidates and 5.8 more months of restricted mean survival over 5 years. Lower mortality was seen across KDPI tiers up to 85%, was consistent across sensitivity analyses and designs, and was seen in both age strata, which matters because 44.1% of RCC candidates were 65 years or older at listing, compared with 20.2% of other candidates. The estimate for KDPI >85% kidneys was imprecise.

The two analytic approaches answer complementary questions. The time-dependent Cox model compares mortality during time spent waiting with mortality after transplantation. The calendar-aligned trials start on each recipient’s transplant date and compare survival with that of candidates who were still waiting that day and could receive a transplant later. Because controls in each trial faced the same calendar period and allocation rules as the recipient, this design compares candidates under the same allocation rules. The analysis aligned on time since listing gave a 14.0-point difference. The waiting group included candidates who received a transplant later in follow-up.

These findings place candidates with RCC-related kidney failure within the established evidence on transplantation. Wolfe et al. reported lower long-term mortality after deceased-donor transplantation than with continued waiting, after an early period of excess risk,^4^ and the day on which cumulative survival became equal in our RCC cohort (day 240) falls within the 5 to 673 days reported across subgroups in that study. Early operative risk could offset some of the survival advantage after transplantation, but this analysis did not test that explanation. Benefit has also been shown for expanded criteria and high-KDPI kidneys and for older candidates,^11,12,19^ who make up a large share of our RCC cohort. Hao et al. reported lower risk of death or deterioration among transplanted than still-waiting candidates with RCC-induced kidney failure,^13^ but compared groups defined by eventual transplant status. Our time-dependent and trial-emulation designs avoid immortal time, separate donor quality and benchmark the RCC estimates against 598,345 other candidates. RCC risk after transplantation remains a concern,^5,20^ yet a systematic review found similar recurrence and survival after treated RCC with transplantation and with dialysis,^8^ recipients with pretransplant cancer, including kidney cancer, had overall survival similar to that of other recipients in the Norwegian national registry,^21^ and in a separately defined registry cohort of 743 deceased-donor recipients with RCC as the diagnosis at transplant, 40 of 273 deaths (14.7%) were attributed to cancer (Table S9).

For urologists who follow patients after nephrectomy, these data support three practical points. First, once oncologic criteria for listing are met,^6,7^ early referral matters, because time on dialysis is associated with worse post-transplant survival^22^ and many older candidates die before a deceased-donor kidney becomes available.^23^ Coordination between the treating urologist and the transplant center about the required cancer-free interval allows listing as soon as that interval is complete. Second, the association between receiving a KDPI ≤85% kidney and survival was similar in candidates with RCC-related kidney failure and other candidates. This estimate can inform counseling once a patient has been listed. Living-donor transplantation was associated with the lowest HR in RCC candidates (0.39), as it was in other candidates. Many viable kidneys are declined before being transplanted, and discarded kidneys overlap substantially in quality with transplanted ones,^24,25^ so declining may carry a real cost. Third, offers of KDPI >85% kidneys deserve individual discussion: in RCC candidates, mortality was higher than with waiting during the first 6 months, but the later estimate was inconclusive. With only 38 recipients and 12 post-transplant deaths, the overall estimate was too imprecise to establish either a survival benefit or harm. These analyses did not establish what drove the early excess mortality. The interaction for KDPI >85% (P = 0.038) rests on 38 recipients and 12 deaths among several transplant states compared, so it is best read as a signal for caution. In the general population, higher-KDPI kidneys benefit older candidates facing long waits,^12^ and the same may hold for selected RCC candidates.

Strengths of this study include national data with death ascertainment that continued after removal from the list and after transplantation, a time-varying exposure that avoids immortal time bias, a calendar-aligned trial design that compares recipients on their transplant date with candidates who were still waiting that day, overlap weighting that achieved exact mean balance of the measured covariates, a direct comparison with the full reference cohort, and independent recomputation of every headline estimate. Estimates were stable across six sensitivity analyses that addressed selection at listing, allocation era and competing transplant pathways.

This study has limitations. The registry does not record tumor stage, histology, grade, margin status, or the cancer-free interval, and RCC was defined by the coded cause of kidney failure at listing, so candidates with treated RCC listed under another diagnosis fall in the reference group. Listed candidates are a selected group that has met cancer-free criteria. Covariates were measured at listing. The registry extract did not contain offer decisions, so the analysis compared recipients with candidates who continued to wait. Residual confounding by health status cannot be excluded, although an unmeasured confounder would need associations of 2.54-fold with both transplantation and death to explain the primary estimate. Only 38 RCC candidates received a KDPI >85% kidney, and post-transplant cancer outcomes come from a separately defined recipient cohort. Death was the only outcome; quality of life and cancer recurrence among waitlisted candidates were not assessed.

## Conclusions

Among waitlisted candidates with kidney failure attributed to RCC, receiving a KDPI ≤85% deceased-donor kidney was associated with longer survival than continued waiting, corresponding to about 16 additional survivors per 100 candidates at 5 years. The relative association was similar in candidates listed for other causes of kidney failure. The estimate for KDPI >85% kidneys was imprecise.

## Author contributions

Junho Song led the study and prepared the manuscript. Esther Kim and Amar Kassim Amar revised the manuscript. Julian Chavarriaga Soto, Jay D. Raman, and Jaime Herrera-Caceres reviewed the manuscript and provided feedback.

## Data availability

The OPTN/UNOS Standard Transplant Analysis and Research (STAR) files, with data through March 31, 2026, are available to eligible researchers through the OPTN data request process. The analysis code is available from the authors.

## References

1. Huang WC, Levey AS, Serio AM, et al. Chronic kidney disease after nephrectomy in patients with renal cortical tumours: a retrospective cohort study. Lancet Oncol. 2006;7(9):735–740. doi:10.1016/S1470-2045(06)70803-8

2. Lane BR, Demirjian S, Derweesh IH, et al. Survival and functional stability in chronic kidney disease due to surgical removal of nephrons: importance of the new baseline glomerular filtration rate. Eur Urol. 2015;68(6):996–1003. doi:10.1016/j.eururo.2015.04.043

3. Campbell SC, Clark PE, Chang SS, Karam JA, Souter L, Uzzo RG. Renal mass and localized renal cancer: evaluation, management, and follow-up: AUA guideline: part I. J Urol. 2021;206(2):199–208. doi:10.1097/JU.0000000000001911

4. Wolfe RA, Ashby VB, Milford EL, et al. Comparison of mortality in all patients on dialysis, patients on dialysis awaiting transplantation, and recipients of a first cadaveric transplant. N Engl J Med. 1999;341(23):1725–1730. doi:10.1056/NEJM199912023412303

5. Karami S, Yanik EL, Moore LE, et al. Risk of renal cell carcinoma among kidney transplant recipients in the United States. Am J Transplant. 2016;16(12):3479–3489. doi:10.1111/ajt.13862

6. Al-Adra DP, Hammel L, Roberts J, et al. Pretransplant solid organ malignancy and organ transplant candidacy: a consensus expert opinion statement. Am J Transplant. 2021;21(2):460–474. doi:10.1111/ajt.16318

7. Chadban SJ, Ahn C, Axelrod DA, et al. KDIGO clinical practice guideline on the evaluation and management of candidates for kidney transplantation. Transplantation. 2020;104(4S1 Suppl 1):S11-S103. doi:10.1097/TP.0000000000003136

8. Boissier R, Hevia V, Bruins HM, et al. The risk of tumour recurrence in patients undergoing renal transplantation for end-stage renal disease after previous treatment for a urological cancer: a systematic review. Eur Urol. 2018;73(1):94–108. doi:10.1016/j.eururo.2017.07.017

9. Rao PS, Schaubel DE, Guidinger MK, et al. A comprehensive risk quantification score for deceased donor kidneys: the kidney donor risk index. Transplantation. 2009;88(2):231–236. doi:10.1097/TP.0b013e3181ac620b

10. Israni AK, Salkowski N, Gustafson S, et al. New national allocation policy for deceased donor kidneys in the United States and possible effect on patient outcomes. J Am Soc Nephrol. 2014;25(8):1842–1848. doi:10.1681/ASN.2013070784

11. Merion RM, Ashby VB, Wolfe RA, et al. Deceased-donor characteristics and the survival benefit of kidney transplantation. JAMA. 2005;294(21):2726–2733. doi:10.1001/jama.294.21.2726

12. Massie AB, Luo X, Chow EKH, Alejo JL, Desai NM, Segev DL. Survival benefit of primary deceased donor transplantation with high-KDPI kidneys. Am J Transplant. 2014;14(10):2310–2316. doi:10.1111/ajt.12830

13. Hao X, Lai W, Xia X, et al. Transplant or dialysis: what’s the better choice for RCC-induced ESRD patients? A 20-year analysis of OPTN/UNOS data. Front Oncol. 2022;12:955771. doi:10.3389/fonc.2022.955771

14. von Elm E, Altman DG, Egger M, et al. Strengthening the Reporting of Observational Studies in Epidemiology (STROBE) statement: guidelines for reporting observational studies. BMJ. 2007;335(7624):806–808. doi:10.1136/bmj.39335.541782.AD

15. Suissa S. Immortal time bias in pharmaco-epidemiology. Am J Epidemiol. 2008;167(4):492–499. doi:10.1093/aje/kwm324

16. Hernán MA, Robins JM. Using big data to emulate a target trial when a randomized trial is not available. Am J Epidemiol. 2016;183(8):758–764. doi:10.1093/aje/kwv254

17. Thomas LE, Li F, Pencina MJ. Overlap weighting: a propensity score method that mimics attributes of a randomized clinical trial. JAMA. 2020;323(23):2417–2418. doi:10.1001/jama.2020.7819

18. VanderWeele TJ, Ding P. Sensitivity analysis in observational research: introducing the E-value. Ann Intern Med. 2017;167(4):268–274. doi:10.7326/M16-2607

19. Rao PS, Merion RM, Ashby VB, Port FK, Wolfe RA, Kayler LK. Renal transplantation in elderly patients older than 70 years of age: results from the Scientific Registry of Transplant Recipients. Transplantation. 2007;83(8):1069–1074. doi:10.1097/01.tp.0000259621.56861.31

20. Acuna SA, Huang JW, Daly C, Shah PS, Kim SJ, Baxter NN. Outcomes of solid organ transplant recipients with preexisting malignancies in remission: a systematic review and meta-analysis. Transplantation. 2017;101(3):471–481. doi:10.1097/TP.0000000000001192

21. Dahle DO, Grotmol T, Leivestad T, et al. Association between pretransplant cancer and survival in kidney transplant recipients. Transplantation. 2017;101(10):2599–2605. doi:10.1097/TP.0000000000001659

22. Meier-Kriesche HU, Port FK, Ojo AO, et al. Effect of waiting time on renal transplant outcome. Kidney Int. 2000;58(3):1311–1317. doi:10.1046/j.1523-1755.2000.00287.x

23. Schold J, Srinivas TR, Sehgal AR, Meier-Kriesche HU. Half of kidney transplant candidates who are older than 60 years now placed on the waiting list will die before receiving a deceased-donor transplant. Clin J Am Soc Nephrol. 2009;4(7):1239–1245. doi:10.2215/CJN.01280209

24. Husain SA, King KL, Pastan S, et al. Association between declined offers of deceased donor kidney allograft and outcomes in kidney transplant candidates. JAMA Netw Open. 2019;2(8):e1910312. doi:10.1001/jamanetworkopen.2019.10312

25. Mohan S, Chiles MC, Patzer RE, et al. Factors leading to the discard of deceased donor kidneys in the United States. Kidney Int. 2018;94(1):187–198. doi:10.1016/j.kint.2018.02.016

