## Supplementary materials, tables, and figure for "Survival after deceased-donor kidney transplantation versus continued waiting among candidates with kidney failure attributed to renal cell carcinoma"

Contents: Sections 1 through 6; Tables S1 to S10; Figure S1. Abbreviations: CI, confidence interval; DCGF, death-censored graft failure; DDKT, deceased-donor kidney transplantation; HR, hazard ratio; IQR, interquartile range; KAS, Kidney Allocation System; KDPI, kidney donor profile index; LDKT, living-donor kidney transplantation; OPTN, Organ Procurement and Transplantation Network; RCC, renal cell carcinoma; ref, reference level; RMST, restricted mean survival time; SMD, standardized mean difference.

### SDC, Materials and Methods

### Section 1. Cohort construction

The source was the OPTN/United Network for Organ Sharing Standard Transplant Analysis and Research files (release with data through March 31, 2026). One record was kept per adult candidate first listed for a kidney-alone transplant from 2005 to 2025 (600,005 candidates). Candidates with a recorded death date before the listing date (110) or before a recorded transplant date (8) were excluded, leaving 599,887 candidates; RCC candidates were listed at 207 centers. The RCC group (1,542) comprised candidates whose coded cause of kidney failure at listing (DGN_TCR) was 3021, renal cell carcinoma; the reference group (598,345) comprised all other candidates. The death date was the earliest patient-level composite death date across every waitlist and transplant record of the patient, so that deaths after removal from the list and after transplantation were captured. The first transplant after listing (from any registration) defined the transplant state; the KDPI, donor type and multiorgan flag were taken from the registration carrying that transplant. No RCC deceased-donor recipient had a missing KDPI.

### Section 2. Counting-process data and Cox models

Time was measured in days from first listing. Each candidate contributed a waiting interval from day 0 to the first transplant (or to death or censoring) and, if transplanted, a second interval in the transplant state from the transplant day to death or censoring. For models with time-varying effects, the post-transplant interval was split at 183 days (early versus later) or on a fine grid for the spline model; splitting at times without events does not change estimates. The risk set at each event time t comprised intervals with start < t ≤ stop. Deaths on the transplant day were assigned to the transplant state. Cox models used Breslow ties and the Lin-Wei cluster-robust variance with listing center as the cluster. Covariates were categorical with reference levels age 55 to 64 years, female sex, White race, no diabetes, preemptive listing, calculated panel-reactive antibody 0%, blood group O, private payer, normal or minimally limited functional status, no prior kidney transplant and listing before KAS. In the subset listed in the KAS era, KAS served as the reference era level (no pre-KAS listings; exposure HRs are invariant to this choice). The time-varying HR after DDKT was modeled with a restricted cubic spline in log days since transplant (knots 7, 30, 120, 365 and 1,826 days); the equal-risk day is the first day after the early peak on which the spline HR falls below 1, and the equal-survival day is the first day on which the accumulated excess hazard after transplantation (Breslow baseline hazard multiplied by HR minus 1, starting at the median time from listing to DDKT) returns to zero. CIs are percentiles of 2,000 draws from the robust covariance of the spline coefficients. The reference spline curve was estimated in a random 25% sample of the reference cohort.

### Section 3. Target trial specification and calendar-aligned sequential trials

Eligibility: RCC candidates alive, listed and untransplanted on a given calendar date. Treatment strategies: receive a KDPI ≤85% deceased-donor kidney on that date versus continue waiting (later transplantation of any type allowed). Assignment: observed transplant on that date; controls were all other eligible RCC candidates on the same date. Time zero: the calendar date of the transplant. Outcome: death from any cause. Follow-up: to death or March 31, 2026, not censored at later transplantation or removal. Causal contrast: intention-to-treat analogue. Analysis: one trial per KDPI ≤85% transplant (597 trials); in the primary analysis up to 50 controls were sampled per trial (29,824 control rows from 1,441 unique candidates), each weighted by the risk-set size divided by the number sampled. A candidate could serve as a control in many trials and later as a treated individual in one trial.

### Section 4. Propensity model and overlap weights

A logistic model for treatment within the stacked trials included the listing covariates of Section 2, time since listing at trial entry (0 to 1, 1 to 2, 2 to 3, 3 to 5 and >5 years) and allocation era at trial entry. Overlap weights were 1 minus the propensity score for treated and the propensity score for controls, multiplied by the sampling weight. Before weighting the maximum absolute SMD was 0.42; after weighting it was <0.001 (logistic overlap weights balance covariate means exactly). Weighted Kaplan-Meier estimators gave survival curves for each arm; RMST was the area under each curve to 5 years, and the RMST gain is the difference in months.

### Section 5. Bootstrap

CIs for the trial estimates came from a candidate-level nonparametric bootstrap with B = 300 replicates: in each replicate RCC candidates were resampled with replacement, the trials were rebuilt from the resampled candidates, the propensity model was refitted and the weighted survival estimates recomputed; percentile limits were used. The number of candidates needed to accept for one additional 5-year survivor is 1 divided by the 5-year difference, with CI limits obtained by inverting the limits of the difference CI (valid because the whole CI lies above zero).

### Section 6. Independent verification

All headline estimates of the original analysis (Python 3.13 with lifelines, statsmodels and scikit-learn) were recomputed independently in Python 3.10 with NumPy and pandas, without importing original code. The reference-cohort and interaction models were refitted on the full reference cohort (598,345 candidates, no sampling). The sequential trials were rebuilt with all eligible controls per trial instead of 50 sampled controls (119,611 control rows; median risk set 206), with a separate candidate-level bootstrap (B = 300). Tolerances were 0.02 for HRs, 0.3 percentage points for percentages (1.0 point for the trials, because sampled and complete control sets differ), 0.6 months for RMST and exact agreement for counts. All 171 records agreed (Table S10). The Cox routine used for verification was corrected during the work so that intervals starting exactly at an event time are excluded from that risk set; all estimates were rerun after the correction and were unchanged.

**Table S1. Characteristics at listing of candidates with kidney failure attributed to RCC, by eventual transplant status**

| Characteristic | DDKT, KDPI ≤85% | DDKT, KDPI >85% | LDKT | Not transplanted |
|---|---|---|---|---|
| Candidates, n | 597 | 38 | 269 | 638 |
| Age, years, median (IQR) | 62.0 (54.0 to 68.0) | 68.5 (62.5 to 72.0) | 63.0 (55.0 to 69.0) | 64.0 (58.0 to 69.0) |
| Dialysis duration at listing, years, median (IQR) | 1.3 (0.0 to 3.2) | 1.0 (0.0 to 2.5) | 0.5 (0.0 to 1.7) | 0.5 (0.0 to 2.0) |
| Body mass index, kg/m$^2$, median (IQR) | 28.9 (25.3 to 33.0) | 28.7 (26.2 to 33.5) | 28.9 (25.4 to 32.2) | 29.6 (26.0 to 33.2) |
| Missing, n | 3 | 0 | 1 | 2 |
| Age group, years, n (%) | | | | |
| 18 to 44 | 46 (7.7) | 0 (0.0) | 25 (9.3) | 16 (2.5) |
| 45 to 54 | 113 (18.9) | 2 (5.3) | 38 (14.1) | 78 (12.2) |
| 55 to 64 | 203 (34.0) | 9 (23.7) | 92 (34.2) | 240 (37.6) |
| ≥65 | 235 (39.4) | 27 (71.1) | 114 (42.4) | 304 (47.6) |
| Male sex, n (%) | 453 (75.9) | 29 (76.3) | 201 (74.7) | 478 (74.9) |
| Race and ethnicity, n (%) | | | | |
| White | 346 (58.0) | 19 (50.0) | 217 (80.7) | 405 (63.5) |
| Black | 181 (30.3) | 16 (42.1) | 24 (8.9) | 154 (24.1) |
| Hispanic | 55 (9.2) | 2 (5.3) | 21 (7.8) | 55 (8.6) |
| Asian | 11 (1.8) | 1 (2.6) | 4 (1.5) | 18 (2.8) |
| Other | 4 (0.7) | 0 (0.0) | 3 (1.1) | 6 (0.9) |
| Diabetes, n (%) | 113 (18.9) | 6 (15.8) | 50 (18.6) | 177 (27.7) |
| Dialysis duration at listing, n (%) | | | | |
| Preemptive | 154 (25.8) | 14 (36.8) | 106 (39.4) | 254 (39.8) |
| <1 y | 114 (19.1) | 4 (10.5) | 61 (22.7) | 126 (19.7) |
| 1 to 3 y | 168 (28.1) | 13 (34.2) | 82 (30.5) | 173 (27.1) |
| ≥3 y | 161 (27.0) | 7 (18.4) | 20 (7.4) | 85 (13.3) |
| Calculated panel-reactive antibody, %, n (%) | | | | |
| 0 | 405 (67.8) | 15 (39.5) | 182 (67.7) | 358 (56.1) |
| 1 to 79 | 41 (6.9) | 2 (5.3) | 22 (8.2) | 38 (6.0) |
| ≥80 | 12 (2.0) | 0 (0.0) | 2 (0.7) | 7 (1.1) |
| Unknown | 139 (23.3) | 21 (55.3) | 63 (23.4) | 235 (36.8) |
| ABO blood group, n (%) | | | | |
| O | 280 (46.9) | 18 (47.4) | 113 (42.0) | 318 (49.8) |
| A | 227 (38.0) | 16 (42.1) | 111 (41.3) | 211 (33.1) |
| B | 61 (10.2) | 3 (7.9) | 33 (12.3) | 93 (14.6) |
| AB | 29 (4.9) | 1 (2.6) | 12 (4.5) | 16 (2.5) |
| Primary payer, n (%) | | | | |
| Private | 189 (31.7) | 13 (34.2) | 124 (46.1) | 248 (38.9) |

| Characteristic | DDKT, KDPI ≤85% | DDKT, KDPI >85% | LDKT | Not transplanted |
|---|---|---|---|---|
| Public | 405 (67.8) | 24 (63.2) | 142 (52.8) | 386 (60.5) |
| Other or unknown | 3 (0.5) | 1 (2.6) | 3 (1.1) | 4 (0.6) |
| Functional status, n (%) | | | | |
| Normal or minimal limitation | 370 (62.0) | 20 (52.6) | 189 (70.3) | 421 (66.0) |
| Impaired | 213 (35.7) | 18 (47.4) | 70 (26.0) | 189 (29.6) |
| Unknown | 14 (2.3) | 0 (0.0) | 10 (3.7) | 28 (4.4) |
| Prior kidney transplant, n (%) | 3 (0.5) | 0 (0.0) | 2 (0.7) | 1 (0.2) |
| Inactive status at listing, n (%) | 170 (28.5) | 10 (26.3) | 99 (36.8) | 258 (40.4) |
| Listing era, n (%) | | | | |
| Before KAS (before Dec 4, 2014) | 277 (46.4) | 9 (23.7) | 112 (41.6) | 258 (40.4) |
| KAS (Dec 4, 2014 to Mar 14, 2021) | 166 (27.8) | 8 (21.1) | 79 (29.4) | 144 (22.6) |
| Acuity circles (from Mar 15, 2021) | 154 (25.8) | 21 (55.3) | 78 (29.0) | 236 (37.0) |
| **Follow-up** | | | | |
| Deaths, n (%) | 198 (33.2) | 12 (31.6) | 73 (27.1) | 331 (51.9) |
| Person-years | 4,981 | 160 | 2,150 | 2,917 |
| Median potential follow-up, y | 10.3 | 3.8 | 9.2 | 8.6 |

Values are n (%) or median (IQR). Transplant status is the first transplant after listing through March 31, 2026. Potential follow-up is the time from listing to the data end date.

**Table S2. Hazard ratios for death for each transplant state versus continued waiting under alternative exposure definitions, RCC and reference cohorts**

| Model | Exposure vs waiting | RCC HR (95% CI) | Reference HR (95% CI) | Ratio of HRs (95% CI) | P value |
|---|---|---|---|---|---|
| Primary | KDPI ≤85% | 0.51 (0.42 to 0.62) | 0.53 (0.51 to 0.54) | 1.02 (0.86 to 1.20) | 0.86 |
| | KDPI >85% | 1.65 (0.87 to 3.13) | 0.74 (0.70 to 0.77) | 1.92 (1.04 to 3.57) | 0.038 |
| | LDKT | 0.39 (0.30 to 0.50) | 0.38 (0.37 to 0.39) | 1.09 (0.86 to 1.39) | 0.47 |
| KDPI tiers | KDPI 0% to 34% | 0.45 (0.35 to 0.59) | 0.47 (0.46 to 0.49) | 0.98 (0.76 to 1.26) | 0.89 |
| | KDPI 35% to 85% | 0.57 (0.46 to 0.71) | 0.58 (0.57 to 0.60) | 1.02 (0.84 to 1.24) | 0.83 |
| | KDPI >85% | 1.64 (0.86 to 3.13) | 0.74 (0.70 to 0.77) | 1.92 (1.03 to 3.56) | 0.039 |
| Any DDKT | Any KDPI | 0.54 (0.45 to 0.65) | 0.53 (0.51 to 0.55) | 1.05 (0.89 to 1.24) | 0.59 |
| 50% cut point | KDPI <50% | 0.49 (0.39 to 0.62) | not estimated | not estimated | |
| | KDPI ≥50% | 0.64 (0.50 to 0.82) | not estimated | not estimated | |
| Continuous | KDPI, per 10 points | 1.08 (1.01 to 1.16) | not estimated | not estimated | |
| Early vs later | KDPI ≤85%, first 6 months | 0.83 (0.49 to 1.40) | not estimated | not estimated | |
| | KDPI ≤85%, after 6 months | 0.49 (0.40 to 0.59) | not estimated | not estimated | |
| | KDPI >85%, first 6 months | 4.85 (1.93 to 12.21) | not estimated | not estimated | |
| | KDPI >85%, after 6 months | 1.22 (0.59 to 2.51) | not estimated | not estimated | |

Time-dependent Cox models adjusted for characteristics at listing; cluster-robust 95% CIs. Exposures are DDKT states (KDPI categories, per-10-point KDPI among DDKT recipients, or period after transplantation) and LDKT. "not estimated" denotes models fitted in RCC candidates only. Ratio of HRs and P from pooled models with transplant state by RCC interaction terms.

### Table S3. Covariate hazard ratios in the primary model, RCC candidates

| Covariate | Level | HR (95% CI) | P |
|---|---|---|---|
| Age group (ref 55 to 64 years) | 18 to 44 years | 0.42 (0.23 to 0.74) | 0.0031 |
| | 45 to 54 years | 0.64 (0.49 to 0.85) | 0.0017 |
| | ≥65 years | 1.57 (1.28 to 1.92) | <0.001 |
| Male sex (ref female) | Yes | 1.37 (1.13 to 1.67) | 0.0016 |
| Race and ethnicity (ref White) | Asian | 0.41 (0.17 to 0.97) | 0.042 |
| | Black | 0.56 (0.44 to 0.71) | <0.001 |
| | Hispanic | 0.80 (0.59 to 1.09) | 0.15 |
| | Other | 0.52 (0.21 to 1.30) | 0.16 |
| Diabetes (ref none) | Yes | 1.42 (1.17 to 1.73) | <0.001 |
| Dialysis duration at listing (ref preemptive) | 1 to 3 years | 1.65 (1.31 to 2.07) | <0.001 |
| | <1 year | 1.60 (1.30 to 1.98) | <0.001 |
| | ≥3 years | 2.04 (1.49 to 2.79) | <0.001 |
| Calculated panel-reactive antibody (ref 0%) | 1% to 79% | 0.82 (0.52 to 1.31) | 0.41 |
| | ≥80% | 1.48 (0.83 to 2.64) | 0.18 |
| | Unknown | 1.05 (0.87 to 1.27) | 0.64 |
| ABO blood group (ref O) | A | 0.95 (0.80 to 1.14) | 0.61 |
| | AB | 1.25 (0.81 to 1.93) | 0.32 |
| | B | 0.92 (0.70 to 1.21) | 0.56 |
| Primary payer (ref private) | Other or unknown | 1.40 (0.74 to 2.67) | 0.30 |
| | Public | 1.18 (0.96 to 1.44) | 0.11 |
| Functional status (ref normal or minimal limitation) | Impaired | 0.98 (0.81 to 1.17) | 0.80 |
| | Unknown | 1.16 (0.81 to 1.66) | 0.43 |
| Prior kidney transplant (ref none) | Yes | 1.00 (0.25 to 4.12) | 1.00 |
| Listing era (ref before KAS) | KAS | 0.97 (0.80 to 1.18) | 0.79 |
| | Acuity circles | 0.72 (0.51 to 1.03) | 0.076 |

Primary time-dependent Cox model (transplant states KDPI ≤85%, KDPI >85% and LDKT versus waiting); values from the independent refit, which reproduced the original log-likelihood exactly.

**Table S4. Time after deceased-donor transplantation at which risk and cumulative survival equal those of continued waiting**

| Time after DDKT | RCC, day (95% CI) | Reference, day (95% CI) |
|---|---|---|
| Equal instantaneous risk of death (transplant vs waiting) | 29 (11 to 107) | 33 (25 to 45) |
| Equal cumulative survival (transplant vs waiting) | 240 (72 to 590) | 218 (185 to 262) |

From the restricted cubic spline model (Figure S1). The reference estimates were obtained in a random 25% sample of the reference cohort.

### Table S5. Overlap-weighted survival and numbers at risk, calendar-aligned trials

| Arm | 1 year (%) | 3 years (%) | 5 years (%) |
|---|---|---|---|
| Accepted a KDPI ≤85% kidney | 96.9 | 90.0 | 83.1 (79.0 to 86.6) |
| Continued waiting | 91.6 | 78.1 | 67.0 (64.0 to 70.8) |
| Difference, points | 5.4 | 11.9 | 16.0 (11.8 to 20.1) |

| Numbers at risk | Year 0 | Year 1 | Year 2 | Year 3 | Year 4 | Year 5 |
|---|---|---|---|---|---|---|
| Accepted (recipients) | 597 | 539 | 476 | 406 | 347 | 300 |
| Waiting (unique candidates) | 1,441 | 1,296 | 1,144 | 988 | 861 | 727 |
| Waiting (trial control rows) | 29,824 | 25,606 | 21,523 | 17,608 | 14,528 | 12,112 |

### Table S6. Absolute survival differences at 5 years under alternative designs

| Design | 5-year survival, accepted (%) | 5-year survival, waiting (%) | Difference, points (95% CI) | RMST gain, months (95% CI) | Candidates accepting per extra 5-year survivor (95% CI) |
|---|---|---|---|---|---|
| Calendar-aligned trials, 50 sampled controls per trial (primary) | 83.1 (79.0 to 86.6) | 67.0 (64.0 to 70.8) | 16.0 (11.8 to 20.1) | 5.8 (4.4 to 7.4) | 6.2 (5.0 to 8.5) |
| Calendar-aligned trials, all eligible controls (independent rebuild) | 83.1 | 67.1 | 16.0 (11.5 to 20.0) | 5.8 (4.4 to 7.2) | 6.3 (5.0 to 8.7) |
| Waiting-time-aligned trials (alternative design) | not reported | not reported | 14.0 (10.3 to 17.6) | 4.3 (3.1 to 5.6) | 7.2 |

Waiting-time-aligned trials aligned controls on time since listing rather than calendar date (values from the original analysis; survival by arm not reported). The candidates-to-accept value for that design is the reciprocal of the point difference.

### Table S7. Sensitivity analyses of the primary hazard ratio, RCC candidates

| Analysis | Candidates / deaths | KDPI ≤85%, HR (95% CI) | KDPI >85%, HR (95% CI) |
|---|---|---|---|
| Primary analysis | 1,542 / 614 | 0.51 (0.42 to 0.62) | 1.65 (0.87 to 3.13) |
| Listed in KAS era or later | 886 / 177 | 0.39 (0.27 to 0.57) | 2.08 (0.80 to 5.39) |
| Active at listing only | 1,005 / 410 | 0.44 (0.34 to 0.57) | 1.54 (0.78 to 3.01) |
| Age ≥60 years at listing | 983 / 429 | 0.55 (0.44 to 0.69) | 2.19 (1.05 to 4.56) |
| Censoring at LDKT | 1,541 / 541 | 0.51 (0.42 to 0.62) | 1.65 (0.88 to 3.12) |
| Censoring at removal for other reasons | 1,542 / 579 | 0.47 (0.38 to 0.59) | 1.45 (0.75 to 2.79) |
| Censoring at multiorgan transplant | 1,542 / 604 | 0.50 (0.41 to 0.61) | 1.64 (0.86 to 3.10) |

Each row refits the primary model in the indicated subset or with the indicated censoring rule.

### Table S8. Prespecified exploratory analysis by age at listing, RCC candidates

| Age at listing | Candidates | Deaths | KDPI ≤85% recipients | Deaths after KDPI ≤85% DDKT | KDPI ≤85% vs waiting, HR (95% CI) |
|---|---|---|---|---|---|
| <65 years | 862 | 301 | 362 | 105 | 0.44 (0.33 to 0.59) |
| ≥65 years | 680 | 313 | 235 | 93 | 0.54 (0.41 to 0.73) |
| Ratio of HRs, ≥65 vs <65 years | | | | | 1.24 (0.82 to 1.87); P = 0.31 |

Models within each stratum used linear age in place of age group. The ratio of HRs compares the two disjoint strata with a z test of the difference in log HRs.

**Table S9. Descriptive outcomes among deceased-donor recipients with RCC as the diagnosis at transplant, by KDPI tier**

| KDPI tier | Recipients | Deaths | Cancer deaths | Death-censored graft failures | 5-year DCGF, % (95% CI) | 5-year cancer death, % (95% CI) |
|---|---|---|---|---|---|---|
| 0% to 34% | 316 | 113 | 20 | 37 | 7.0 (4.3 to 10.5) | 2.4 (1.0 to 5.0) |
| 35% to 85% | 384 | 145 | 16 | 58 | 12.2 (8.7 to 16.2) | 2.6 (1.1 to 5.0) |
| 85% | 43 | 15 | 4 | 9 | 22.5 (9.4 to 39.0) | 16.5 (4.9 to 34.0) |

Separate cohort of adult kidney-alone deceased-donor recipients transplanted from 2005 to 2025 whose diagnosis at transplant was RCC (n = 743; median observed follow-up 4.4 years). Cancer deaths were deaths with a cancer cause code. Cumulative incidence by Aalen-Johansen with death as a competing event. Values from the original analysis (provenance only; this cohort was not available for independent recomputation).

**Table S10.** Verification of headline estimates

| Quantity | Primary analysis | Independent recomputation | Verdict |
|---|---|---|---|
| RCC HR, DDKT KDPI ≤85% vs waiting | 0.51 | 0.51 | agree |
| RCC HR, DDKT KDPI >85% vs waiting | 1.65 | 1.65 | agree |
| RCC HR, LDKT vs waiting | 0.39 | 0.39 | agree |
| Reference HR, DDKT KDPI ≤85% (full cohort, no sampling) | 0.53 | 0.53 | agree |
| Ratio of HRs, KDPI ≤85%, RCC vs reference | 1.02 | 1.02 | agree |
| Ratio of HRs, KDPI >85%, RCC vs reference | 1.92 | 1.92 | agree |
| 5-year survival difference, points | 16.0 | 16.0 | agree |
| 5-year RMST gain, months | 5.8 | 5.8 | agree |
| E-value, primary HR | 2.54 | 2.54 | agree |

Independent recomputation used all eligible controls for the trials (sampled controls in the primary analysis).

**Figure S1. Time-varying hazard ratio for death after deceased-donor transplantation versus continued waiting, RCC candidates (solid blue line, light blue 95% CI band) and reference cohort (dashed red line, light red band; random 25% sample), from restricted cubic splines in log time since transplantation. Vertical dotted lines mark the day of equal instantaneous risk (day 29) and of equal cumulative survival (day 240) for RCC candidates.**

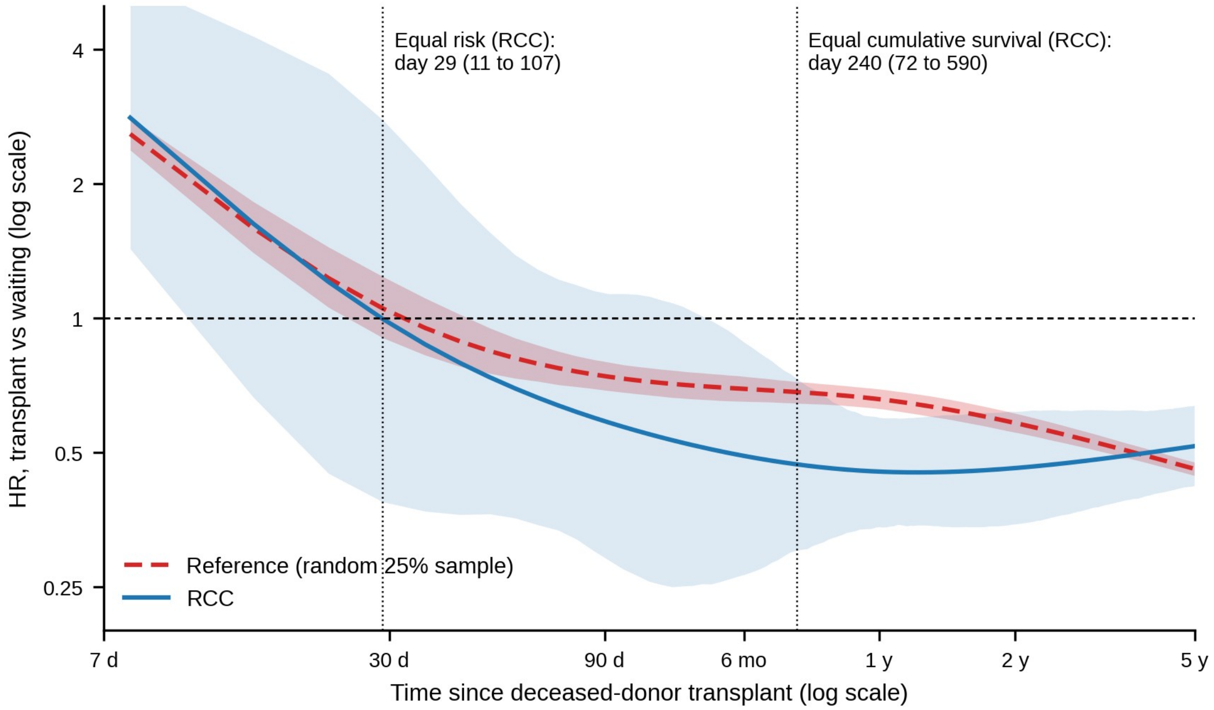
